# Developing and testing an arts-based, digital knowledge translation tool for parents about childhood croup

**DOI:** 10.1101/2021.06.03.21257424

**Authors:** Shannon D. Scott, Anne Le, Lisa Hartling

**Affiliations:** Faculty of Nursing, University of Alberta, Edmonton, Canada; Department of Pediatrics, University of Alberta, Edmonton, Canada

## Abstract

Croup is a common viral illness affecting 80,000 children annually in Canada. Between 7-31% of children seen in an ED for croup are admitted to hospital due to health care provider apprehension. However, over 60% of children with croup experience mild symptoms that can be safely managed at home. Emerging evidence suggests that initiatives targeting healthcare consumers (i.e., patients, parents, families) can inform decision making and shape treatment expectations. Previous research demonstrates that innovative media are superior to traditional standard health sheets for transferring information to consumers.

The purpose of this project was to develop, refine, and test the usability of a whiteboard animation video for parents about childhood croup. Parents rated the tools highly across all usability items, suggesting that creative tools developed using multi-method development processes can help facilitate the uptake of health information in parents.

## Introduction

Croup is a common viral illness affecting 80,000 children annually in Canada and is among the three most common reasons for pre-school aged children to visit an Emergency Department (ED) [1-2]. Between 7-31% of children seen in an ED for croup are admitted to hospital due to health care provider apprehension of the potential for respiratory failure and death, although endotracheal intubation is uncommon (0.4-1.4% of hospitalized children), and death is rare (0.5% of intubated children) [3]. Of note, over 60% of children with croup experience mild symptoms that can be safely managed at home [4].

Emerging evidence suggests that initiatives targeting healthcare consumers (i.e., patients, parents, families) can inform decision making and shape treatment expectations [5-6]. Interventions or tools to communicate complex child health information to parents about childhood croup symptoms, home management strategies, and when to seek medical care, may have the potential to reduce unnecessary ED utilization [7]. However, conventional approaches to provide health information (e.g., standardized written instruction sheets) have been found by patients to be unsatisfactory because they often contain language that is too complex and laden with excessive medical jargon [8-12]. Previous research demonstrates that innovative media are superior to traditional standard health sheets for transferring information to consumers [13-15]. Additionally, interventions or tools incorporating narrative and artistic elements have been shown to generate emotional impact and facilitate efficient memory and retrieval [16-17].

Whiteboard animation videos are a digital medium that utilize narrative and art and encourage interactivity. The main foci of these videos are ‘in progress’ graphic drawings complemented by a voiced narrative. There is increasing interest in using whiteboard animation videos as a health communication tool; however, there is little published health research on their development and usability for parents regarding complex child health topics. The purpose of this study was to develop, refine, and test the usability and uptake of a whiteboard animation video for parents about childhood croup.

## Methods

A multi-method design was used to address the following research question: what is the usability and uptake of a whiteboard animation video for parents about childhood croup? Research ethics approval was obtained from the University of Alberta Health Research Ethics Board [Pro00050107]. Additional ethics and operational approvals were obtained by individual hospitals to conduct usability testing.

### Iterative Development Process

Active parent and stakeholder engagement was a key focus of all stages of the iterative development process of the whiteboard animation video. A timeline of the processes involved is included in **Appendix A**. Previous research by the study team collected and analyzed qualitative data on parental experiences with childhood croup to inform the development of a composite narrative for the whiteboard animation video, complemented by research-based recommendations from the TRanslating Emergency Knowledge for Kids (TREKK) Bottom Line Recommendations (BLR) [16, 18]. The interview guide used to elicit parents’ experiences can be found in **Appendix B**. Results from these interviews are published elsewhere [16]. Updates to the research-based recommendations are completed regularly, with literature searches updated on a bi-annual basis [18]. Through a competitive process, we selected an animation studio with writers, digital animators, and voice actors to develop a storyboard, script, and prototype video. The prototype was shared with two groups of parents with national representation: 1) the TREKK Parent Advisory Group, 2) the Canadian Family Advisory Network. The same prototype was also shared with emergency medicine clinicians at the annual Pediatric Emergency Research Canada (PERC) research meeting. Feedback was solicited from these stakeholders and used to refine the video content, script, and visuals.

The video was 2 minutes and 56 seconds, narrated in the third person, and portrayed a single mother and her young male child’s experience with croup. The story is set around bedtime when the child suddenly starts exhibiting a cough. The video includes a sound clip of a croup cough, which is often distinct from other types of cough. The narrator describes what croup is and provides measures parents could take to manage a mild case of croup. The video also provided parents with information about what is likely to happen if they were to go to the hospital for croup symptoms. Finally, the video also includes information about preventative measures parents could take against croup, including washing hands and cleaning surfaces (**Appendix C**).

Usability refers to the capacity of a tool or intervention to be used easily, efficiently, and satisfactorily by people [19]. Offering interventions or tools in a digital format enables more people to benefit from the intervention, however ensuring the digital intervention is usable is critical prior to widespread dissemination [20-22]. Therefore, this study employed a multi-method approach.

### Surveys

Parents were recruited to participate in an electronic survey in three ED waiting rooms across Canada representing urban (Stollery Children’s Hospital), rural (Portage District General Hospital), and remote (Stanton Hospital) health regions. Members of the study team approached parents in the ED to determine interest and study eligibility. Study team members were available in the waiting rooms to provide technical assistance and answer questions as parents were completing the surveys.

Survey content was informed by a systematic search of over 180 usability evaluations [19]. The survey was comprised of 8, 5-point Likert items assessing: 1) usefulness, 2) simplicity, 3) level of engagement, 4) satisfaction, 5) quality of information, and 6) perceived value (**Appendix D**). Two free text boxes were also included.

Data were collected using the FluidSurveys platform on iPads. This platform allows for synchronous and asynchronous data collection and data is stored on a secure, Canadian server [23]. As previously reported, iPads were optimized with rigorous security features, including passcode login, data encryption, GPS tracking, and remote wipe capability [24]. Survey data was cleaned and analysed using SPSS v. 24 [25]. Descriptive statistics and measures of central tendency were generated. Survey data from the free text boxes was analyzed using content analysis.

### Focus Groups

Parents were purposively recruited to participate in one focus group in each of the three regions. Focus groups are an efficient, cost-effective data collection method that provides opportunities to generate rich data while also observing group dynamics, and levels of consensus on topics [26-27]. In two regions, parents were recruited from existing parent advisory groups involved in child health research. In the third region, the local public health unit distributed flyers about the focus group to their parent groups.

The semi-structured focus group guide was comprised of questions about: a) video viewing experience, b) video attributes that were useful, c) video elements that were not helpful, d) perceptions of the video’s utility, and e) recommended revisions (**Appendix E**). Each aspect of the video (i.e., narrative, visual appeal, health information, engagement and interactivity) was explored. Focus groups were audio recorded on iPads with the same security features. Digital recordings were securely transferred to a third party for verbatim transcription. Focus group transcripts were cleaned and analyzed using NVivo 11 [28]. Data was analyzed deductively using the semi-structured interview guide to develop broad categories. Focus group data was then coded, codes were placed into the broad categories, and the categories synthesized into themes. Findings from the focus groups were used to inform final revision of the tool prototypes.

## Results

### Survey Results

Thirty-eight surveys were completed by parents. Participant demographics are presented in Table 1. The results of the usability survey measures are presented in Table 2. Participants gave favourable scores for all 8 usability questions, especially for “it is simple to use” and “I am satisfied with it”. Further analyses were conducted to compare responses based on region (Urban vs. Rural vs. Remote and Urban vs. Non Urban). Results did not show significant differences between the sites.

**Table 1.** Demographic characteristics of parents who assessed the usability of the whiteboard animation video (N=38)

| Characteristic | n (%) |
| --- | --- |
| Sex |  |
| Female | 30 (78.9) |
| Male | 8 (21.1) |
| Age |  |
| 20-29 years | 4 (10.5) |
| 30-39 years | 18 (47.3) |
| 40-49 years | 10 (26.3) |
| 50-59 years | 3 (7.9) |
| 60 years and older | 3 (7.9) |
| Marital Status |  |
| Married/Partnered | 28 (73.7) |
| Single/Separated/Divorced/Widowed | 10 (26.3) |
| Education |  |
| Primary school | 2 (5.3) |
| Some high school | 2 (5.3) |
| High school diploma | 8 (21.1) |
| Some post-secondary | 4 (10.5) |
| Post-secondary certificate/diploma | 12 (31.6) |
| Post-secondary degree | 5 (13.2) |
| Graduate degree | 5 (13.2) |
| Household Income |  |
| Less than \$25,000 | 4 (10.5) |
| \$25,000-\$49,000 | 8 (21.1) |
| \$50,000-\$74,000 | 4 (10.5) |
| \$75,000-\$99,000 | 8 (21.1) |
| \$100,000-\$149,000 | 4 (10.5) |
| \$150,000 and over | 7 (18.4) |
| Prefer not to answer | 3 (7.9) |
| Child Ages |  |
| Under 1 year | 5 (13.2) |
| 1-2 years | 10 (26.3) |
| 3-5 years | 12 (31.6) |
| 6-8 years | 9 (23.7) |
| 9-11 years | 10 (26.3) |
| 12-15 years | 9 (23.7) |
| 15-17 years | 8 (21.1) |
| 18 years and older | 8 (21.1) |
| Research Site |  |
| Urban | 25 (65.8) |
| Rural | 6 (15.8) |
| Remote | 7 (18.4) |

**Table 2.** Means of participant responses to the usability survey

| Usability Measures | Mean (SD) |
| --- | --- |
| It is useful. | 4.61 (0.63) |
| It meets my information needs. | 4.50 (0.63) |
| It is simple to use. | 4.86 (0.35) |
| I can use it without written instructions or additional help. | 4.42 (0.50) |
| It is fun to use. | 4.08 (0.50) |
| I am satisfied with it. | 4.84 (0.51) |
| I would use it in the future. | 4.41 (0.76) |
| I would recommend it to a friend. | 4.47 (0.59) |

The survey contained two free-text questions asking participants to list the most positive and negative aspects of the video. Thirty-four participants provided feedback on positive aspects of the video and their responses were grouped into four main categories: informative, easy to understand, short and to the point, and visual appeal (n=4). Fifteen participants provided feedback on negative aspects of the video, which were grouped into four main categories: narrator spoke too fast, too wordy, too long, and the moving hand drawing the content was distracting.

### Focus Group Findings

Nineteen parents participated in 3 focus groups; one focus group in each of the three regions. The average number of participants was six per focus group. Participant demographics are presented in Table 3. Focus group length ranged from 23 minutes to 42 minutes.

**Table 3.** Demographic characteristics of parents who participated in focus group discussions (N=19)

| Characteristic | n (%) |
| --- | --- |
| Sex |  |
| Female | 11 (57.9) |
| Male | 8 (42.1) |
| Age |  |
| 20-29 years | 3 (15.8) |
| 30-39 years | 4 (21.1) |
| 40-49 years | 10 (52.6) |
| 50-59 years | 1 (5.3) |
| 60 years and older | 1 (5.3) |
| Marital Status |  |
| Married/Partnered | 13 (68.4) |
| Single/Separated/Divorced/Widowed | 5 (31.6) |
| Education |  |
| Primary school | 0 (0.0) |
| Some high school | 0 (0.0) |
| High school diploma | 1 (5.3) |
| Some post-secondary | 1 (5.3) |
| Post-secondary certificate/diploma | 3 (15.8) |
| Post-secondary degree | 10 (52.6) |
| Graduate degree | 2 (21.1) |
| Household Income |  |
| Less than \$25,000 | 3 (15.8) |
| \$25,000-\$49,000 | 2 (10.5) |
| \$50,000-\$74,000 | 2 (10.5) |
| \$75,000-\$99,000 | 2 (10.5) |
| \$100,000-\$149,000 | 3 (15.8) |
| \$150,000 and over | 5 (26.3) |
| Prefer not to answer | 2 (10.5) |
| Number of Children |  |
| 1 | 4 (21.1) |
| 2 | 7 (36.8) |
| 3 | 3 (15.8) |
| 4 | 4 (21.1) |
| Research Site |  |
| Urban | 8 (42.1) |
| Rural | 4 (21.1) |
| Remote | 7 (36.8) |

Participants enjoyed the whiteboard animation medium. In particular, the parents liked that the video about croup was succinct, well-paced, and had a realistic and informative storyline. In addition to the helpful information provided, the video resonated with parents: *“I really liked [the croup video] because it brought back my own memories and the stories and what that felt like. So it would give you an emotional response, which was great, because I can imagine at that time, how that would be a very good video to watch” (Participant 7, focus group 2)*.

While the short, succinct nature of the video was largely described as an asset, some parents did want more information on the cause of the illness, how to contact health providers, and how to prevent future bouts of croup: *“What I liked about [the croup video] was how to avoid it in the future […] ‘Cause I think prevention’s just as important as… you know, what to do when it’s acute and you have to get to the Emerg*.*” (Participant 1, focus group 1)*. They also identified issues of representation, such as the importance of including diversity (i.e., race, gender, language, etc.), considering social-demographic factors in the video narrative: *“The baby monitor is irrelevant, whether you hear you baby coughing through a baby monitor or not. And some families don’t have baby monitors. And driving to the hospital […]. That’s irrelevant. It’s - get them to the hospital*.*” (Participant 1, focus group 2)*. Participants suggested translating the video and adding sub-titles to reinforce the audio content and make the video accessible to those with hearing impairments: *“What I’m seeing online is that they’re putting subtitles on – for everything, for people who are, you know, hard of hearing or for people who just don’t want their volume on – which if you have a sleeping baby, sometimes they don’t want the volume on, right?” (Participant 6, focus group 3)*. Parents also indicated that delivering this information at the appropriate time is an important consideration and the timing might not always support the use of a video: *“Well, it depends how I’m gonna access the information and when I need it. So… you know, when somebody hands me something in the Emergency Room, I’m probably gonna refer to it later and ask the questions of the person who’s standing in front of me. But I like the videos very much for, if I have questions and I Google, “croup” and the video shows up, in less than three minutes I get some good information about, ‘don’t panic - this is what’s going on. Here’s how you determine if you need to go any further’*” *(Participant 1, focus group 1)*.

## Conclusions

Fundamental to meaningful parent involvement in child health decision making is ensuring that they have access to engaging, understandable, and easily available information. Innovative digital media using narrative and artistic elements is a promising approach for communicating complex health information to parents and families.

The results of this study demonstrate that a digital whiteboard animation video for parents’ is highly usable, well accepted, and has excellent uptake. More research is needed to determine the effectiveness of digital knowledge translation tools for parents on a variety of health conditions and health outcomes.

**The tools can be found here: http://www.echokt.ca/tools/croup/**

Note: Our KT tools are assessed for alignment with current, best-available evidence every two years. If recommendations have changed, appropriate modifications are made to our tools to ensure that they are up-to-date.

## Data Availability

The data referred to in the manuscript is stored at the University of Alberta and under Research Ethics Board guidelines, are not available for access.

## Other Outputs from this Project

## Awards

Scott, S.D., Knisley, L., Klassen, T., Hartling, L., Archibald, M., Albrecht, L. Hamm, M. (2015). A barky, seal-like cough in children: An innovative tool for parents that merges the best research evidence with the power of art and story. Prize: IHDCYH Talks, CIHR ($3,000).

## Presentations & Research Conferences

Scott, S.D., Albrecht, L., Knisley, L., Klassen, T., Hartling, L. Usability results of arts-based KT tools for pediatric acute gastroenteritis and croup (oral presentation). KT Canada Annual Scientific Meeting, Vancouver, BC. June 7-8, 2018.

Scott, S.D., Albrecht, L., Knisley, L., Klassen, T., Hartling, L. (2017). Usability results of art-based knowledge translation tools for pediatric acute gastroenteritis and croup. 31st Margaret Scott Wright Research Day. Edmonton, Alberta. November 3, 2017.

Albrecht, L., Schreiber, S., Scott, S.D., Hartling, L. Living systematic reviews for up-todate evidence: case studies on pediatric croup and acute gastroenteritis. 24th Cochrane Colloquium, Seoul, Korea. Oct 23-27, 2016

Scott, S.D., Hartling, L., Albrecht, L., Archibald, M., Hamm, M., Knisley, L. & Klassen, T.P. (2014). Knowledge translation tools for parents with children with croup and gastroenteritis [Poster presentation]. Presented at Women & Children’s Health Research Institute Annual Research Day, Edmonton, AB. November 12, 2014.

Scott, S.D., Hartling, L., Albrecht, L., Archibald, M., Hamm, M., Knisley, L. & Klassen, T.P. (2014). Knowledge translation tools for parents with children with croup and gastroenteritis [Poster presentation]. Presented at 28th Annual Margaret Scott Wright: Research and Innovation Day, Edmonton, AB. November 7, 2014.

## Related Papers

Hartling, L., Scott, S.D., Johnson, D., Bishop, T. & Klassen, T. (2013). A randomized controlled trial of storytelling as a communication tool. *PLoS One*, 8(10): e77800. doi: 10.1371/journal.pone.0077800

Scott, S.D., Hartling, L., O’Leary, K., Archibald, M. & Klassen, T. (2012). Stories - A novel approach to transfer complex health information to parents: A qualitative study. *Arts & Health: An International Journal for Research, Policy & Practice*, 4(2), 162-173. doi: 10.1080/17533015.2012.656203

Hartling, L., Scott, S.D., Pandya, R., Johnson, D., Bishop, T., & Klassen, T.P. (2010). Storytelling as a communication tool for health consumers: Development of an intervention for parents of children with croup. Stories to communicate health information. *BMC Pediatrics*, 65: 1-10. doi:10.1186/1471-2431-10-64

## Related Tools

Hartling, L. & Scott, S.D. (2006). Things We Take for Granted - Severe Croup. University of Alberta.

Hartling, L. & Scott, S.D. (2006). A Late Night Trip to the ED - Moderate Croup. University of Alberta.

Hartling, L. & Scott, S.D. (2006). Managing Croup at Home - Mild Croup. University of Alberta.

## Appendices

## Appendix A Project Timeline

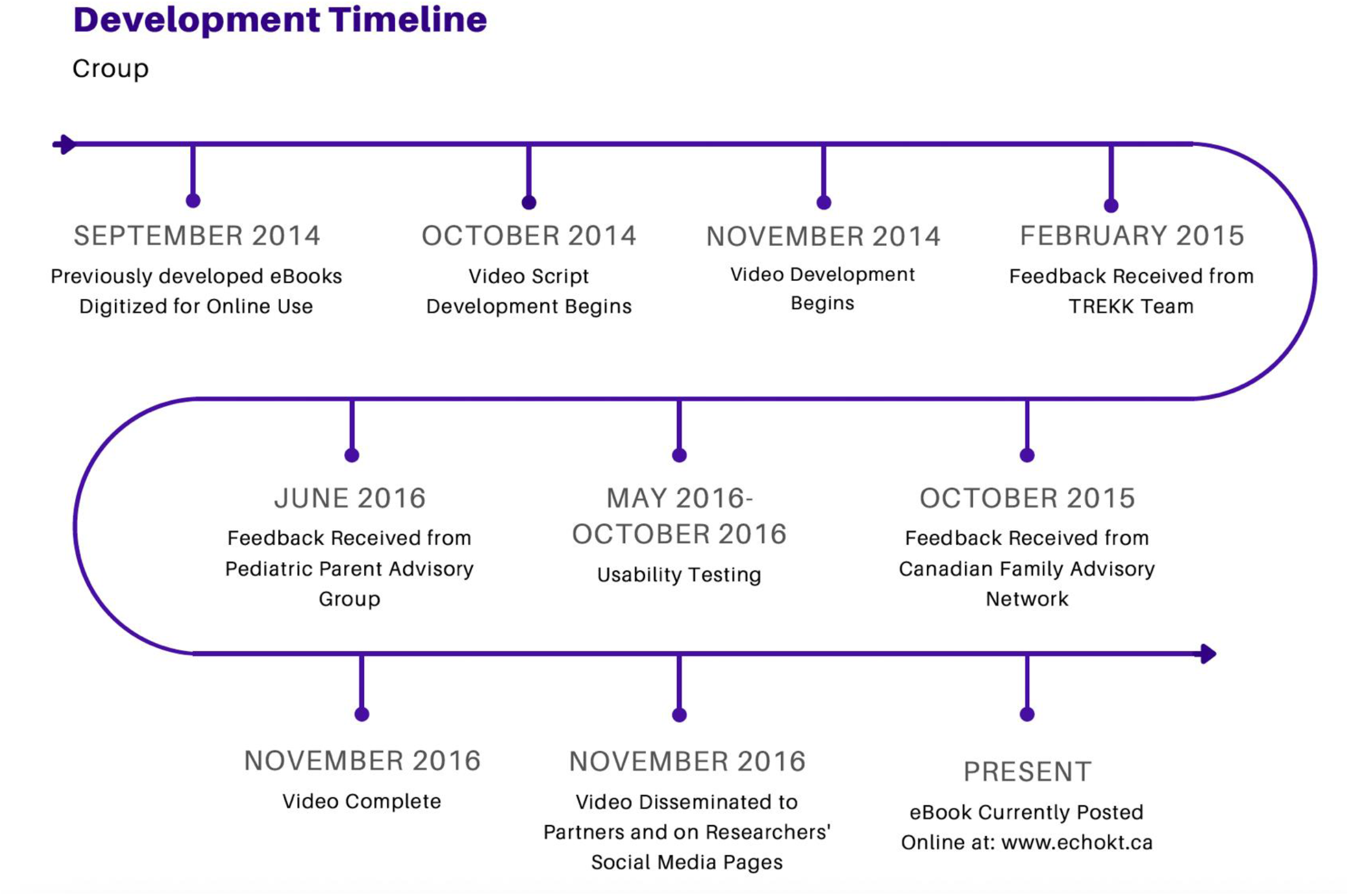

## Appendix B Qualitative Interview Guide

Hello, this is. I met you at the Stollery Children’s Hospital. I was interviewing parents in the Emergency Department for a Storytelling project. You agreed to participate in the study and were given an information letter and consent form to sign at the hospital. Do you remember? I mentioned that I would be contacting you in 10 days. I would like to ask you several questions regarding your experience with your child’s illness once leaving the hospital. Is this a good time to talk? The interview will talk about 15 minutes.

You do not have to participate in this interview if you do not wish to. You do not have to answer any question you are not comfortable answering. You have the right to withdraw at any time.

Do we have your consent to conduct this telephone interview?

Do you have any questions before I begin?

Let’s start off with a broad question;

1. Could you please tell me about your experience at the Stollery Emergency Department? ** Probe for both feelings and more technical/medical experiences <u>Feeling prompts</u>
  ▪ Child/family anxiety levels
  ▪ Emotional response
  ▪ What were you most concerned with while at the ER?
  ▪ Full impact on family
  ▪ Follow-up research/advice received
  ▪ Difference between being in the emergency department and being home
  <u>Medical prompts</u>
  ▪ Medications administered/discharge needs
  ▪ Interaction with staff
  ▪ Logistical problems associated with food/parking/childcare etc.
  ▪ Child’s response to ER visit
2. How has your child’s health been since the trip to the emergency? <u>prompts</u>
  ▪ What on-going treatments were required?
  ▪ Had they been to see a family doctor and/or returned to ED in the past 10 days?
  ▪ Resolution of symptom time and current status
3. Did you receive any information about your child’s illness prior to leaving the hospital? <u>prompts</u>
  ▪ Did parent/guardian ask questions?
  ▪ Use the Internet?
  ▪ Get information sheets from nurses?
  ▪ If they received storytelling booklet, where is it now?
4. Was this information helpful?
5. How do you feel about the overall experience at the Stollery ER and your experience having an ill child?
6. What could have been done differently to make it easier for you during your visit to the ER?

Thank you for your thoughtful answers. This is the end of the interview. Do you have any additional comments?

## Appendix C Croup Video

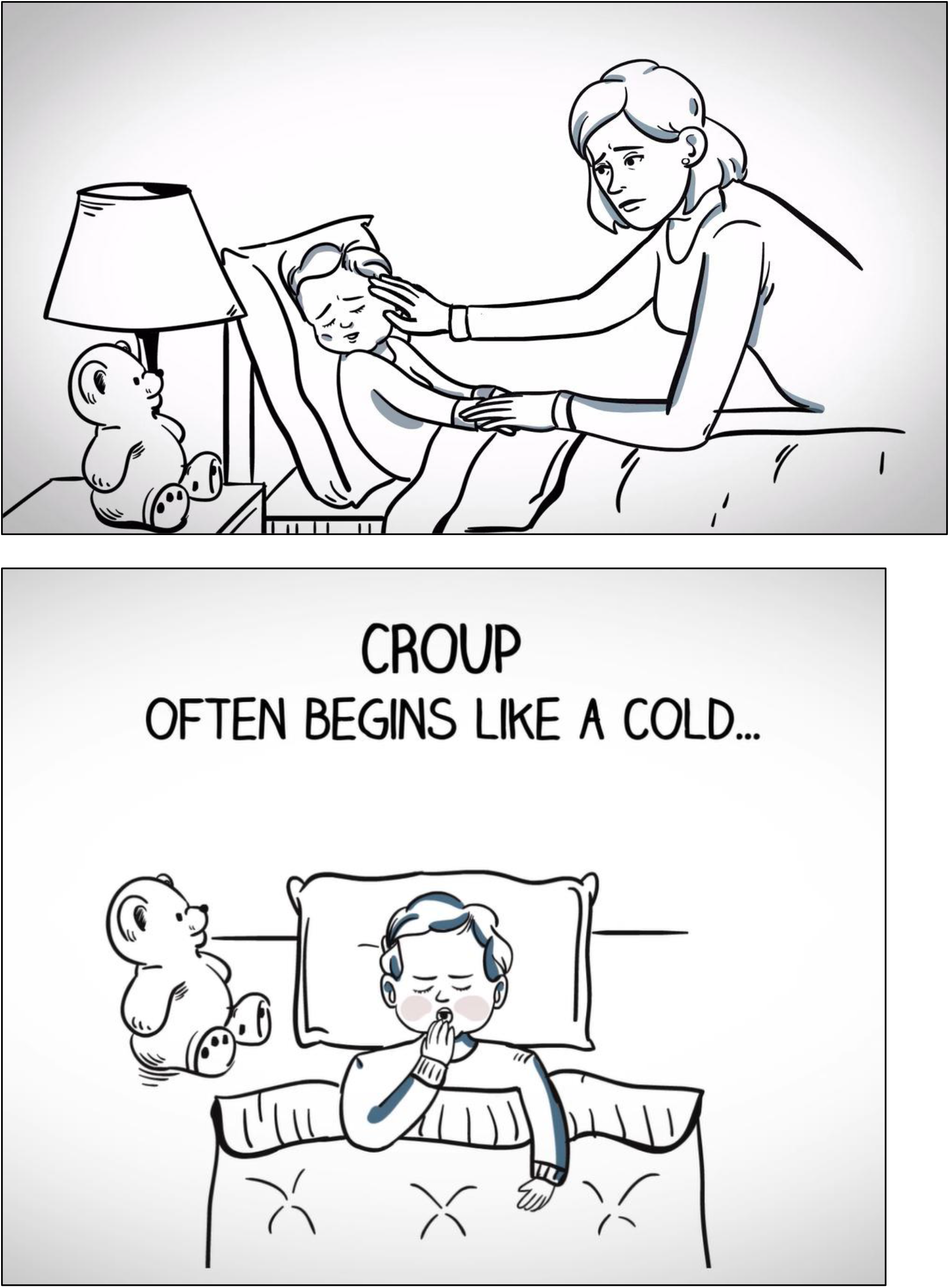
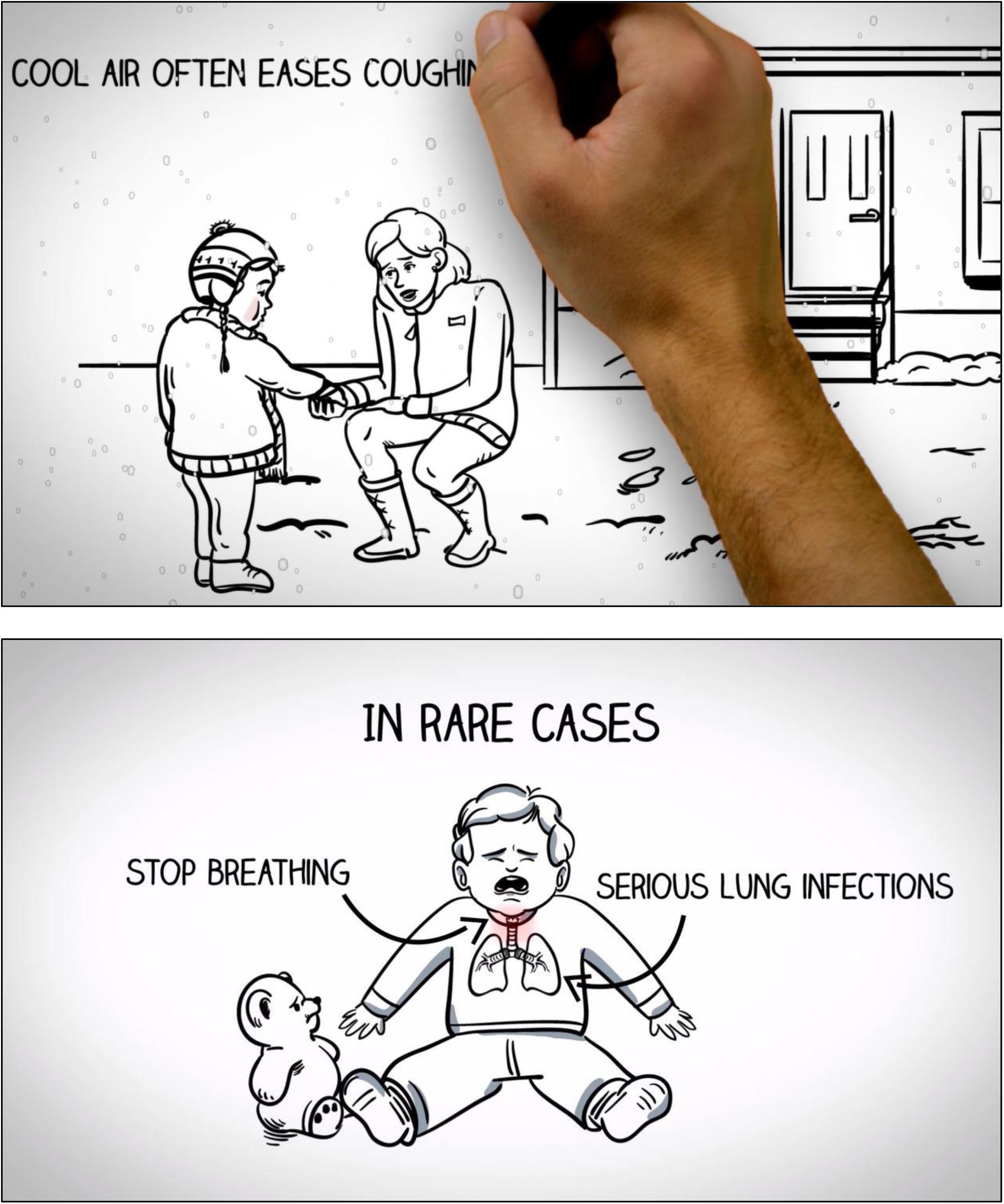
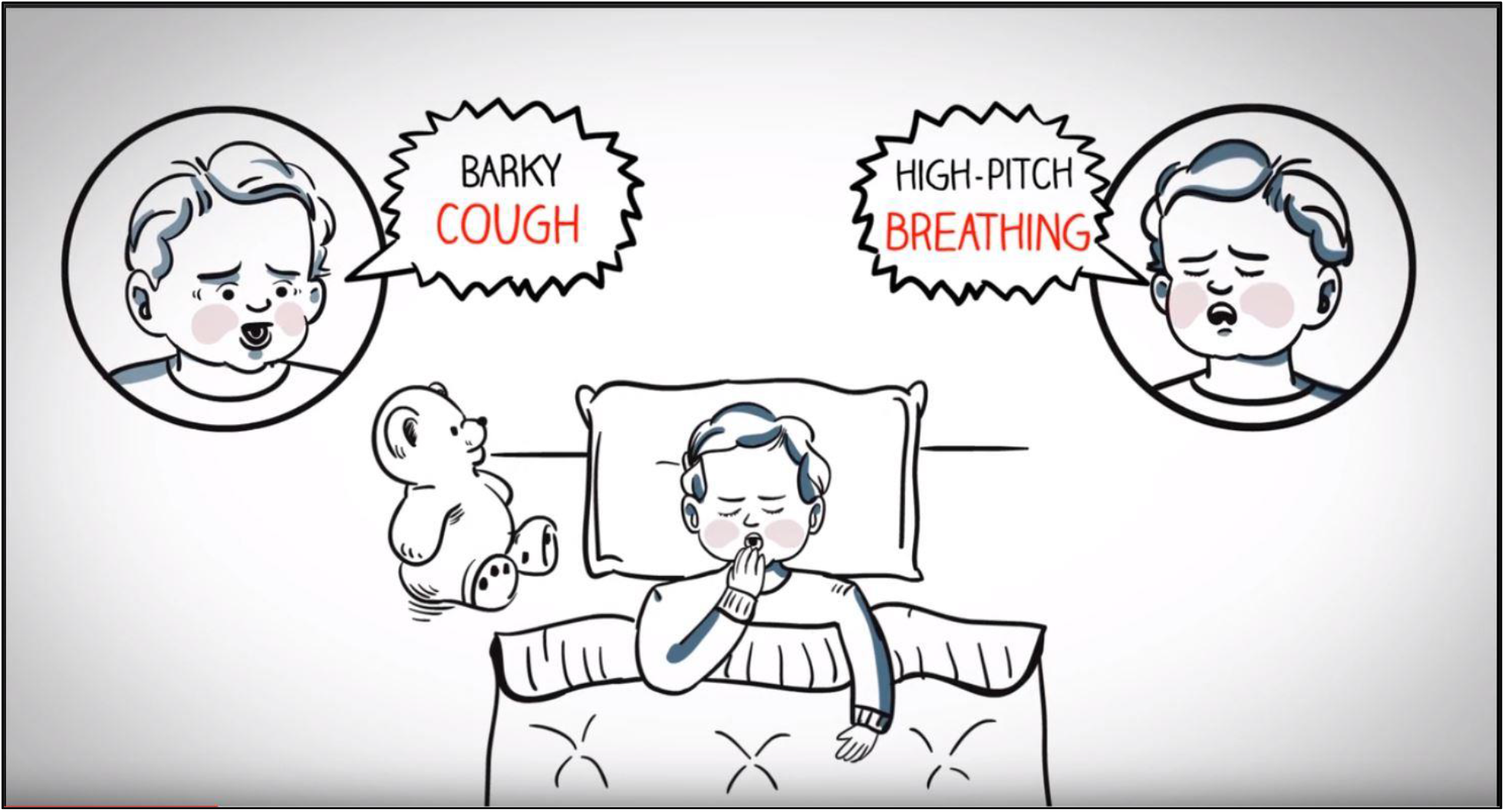

## Appendix D Usability Testing Survey

SECTION 1: Demographics

1. What is your gender?
  □ Male
  □ Female
2. What is your Age?
  Less than 20 years old
  □ 20-30 years
  □ 31-40 years
  □ 41-50 years
  □ 51 years and older
3. What is your Marital Status?
  □ Married
  □ Single
4. What is your gross annual household income?
  □ Less than $25,000
  □ $25,000-$49,999
  □ $50,000-$74,999
  □ $75,000-$99,999
  □ $100,000-$149,999
  □ $150,000 and over
5. What is your highest level of education?
  □ Some high school
  □ High school diploma
  □ Some post-secondary
  □ Post-secondary certificate/diploma
  □ Post-secondary degree
  □ Graduate degree
  □ Other
6. How many children do you have? _______
7. How old are your children? _______________

SECTION 2: Assessment of attributes of the arts-based, digital tools

***\*participant is randomized to view 1 of 2 digital tools then automatically directed to the survey***

1. It is useful. [5-point Likert Scale]
2. It meets my information needs. [5-point Likert Scale]
3. It is simple to use. [5-point Likert Scale]
4. I can use it without written instructions or additional help. [5-point Likert Scale]
5. It is fun to use. [5-point Likert Scale]
6. I am satisfied with it. [5-point Likert Scale]
7. I would use it in the future. [5-point Likert Scale]
8. I would recommend it to a friend. [5-point Likert Scale]
9. List the most negative aspects: [open text]
10. List the most positive aspects: [open text]

## Appendix E Focus Group Guide

Good morning/afternoon. Thank you for taking the time to meet with us. We would like to ask you several questions about your impressions of the eBooks and Whiteboards. Our conversation is being transcribed to ensure that we have an accurate summary of your opinions. All the information we collect will be kept confidential. You may refuse to answer any questions or leave the focus group at any time. Do you have any questions before we begin? Please feel free to ask questions at any time during the interview.

Let’s get started;

1. Tell us your <u>first impressions</u> of the arts-based digital tools provided to you in advance of this focus group.
  - eBooks
  - Whiteboards
  - Describe the <u>usability</u> of these digital tools
2. Did the tools provide you with useful information about pediatric croup OR pediatric gastroenteritis?
  - Explain how
  - Tell me what information you learned.
  - If yes, what information was most useful? Least useful?
  - If no, what information was missing or inadequate?
3. What <u>new</u> information did you learn?
  - Difference between eBooks and Whiteboards
4. How do you anticipate that this information will influence your experience in the future?
5. How will the information provided in the digital tools help you make decisions?
  - Difference between eBooks and Whiteboards
6. How did accessing information in this format (eBooks and Whiteboards) compare with the more traditional parent educational sheets? What digital tool did you prefer?
7. Overall, what is your impression of the eBook? Why did you like it? Why did you dislike? <u>prompt</u>
  ▪ Story/narrative
  ▪ Art
  ▪ Aesthetics/Style
  ▪ Size
  ▪ Color
  ▪ Format
  ▪ Readability
  ▪ Interactivity/Engagement
  ▪ Embedded audio
  ▪ Length
8. Overall, what is your impression of the whiteboard? Why did you like it? Why did you dislike? <u>prompt</u>
  ▪ Drawing style
  ▪ Story/Narrative
  ▪ Size
  ▪ Color
  ▪ Format
  ▪ Voice of the narrator
  ▪ Length
  ▪ Interactivity/Engagement
9. Are there any recommendations for additions, changes to the digital tools?
10. Would you recommend this digital knowledge translation tools to other parents?

Thank you for your thoughtful responses to our questions. Are there any other comments/concerns about the digital tools that have not yet touched upon?

